# Comparing reactions to COVID-19 and influenza vaccinations: data from patient self-reporting, smartwatches and electronic health records

**DOI:** 10.1101/2023.06.28.23292007

**Authors:** Matan Yechezkel, Gary Qian, Yosi Levi, Nadav Davidovitch, Erez Shmueli, Dan Yamin, Margaret L. Brandeau

## Abstract

**Background:** Public reluctance to receive COVID-19 vaccination is due in large part to safety concerns. We compare the safety profile of the BNT162b2 COVID-19 booster vaccine to that of the seasonal influenza vaccine, which has been administered for decades with a solid safety record and a high level of public acceptance.

**Methods:** We study a prospective cohort of 5,079 participants in Israel (the PerMed study) and a retrospective cohort of 250,000 members of Maccabi Healthcare Services. We examine reactions to BNT162b2 (Pfizer-BioNTech) mRNA COVID-19 booster vaccinations and to influenza vaccination. All prospective cohort participants wore a Garmin Vivosmart 4 smartwatch and completed a daily questionnaire via smartphone. For the prospective cohort, we compare pre-vaccination (baseline) and post-vaccination smartwatch heart rate data and a stress measure based on heart rate variability, and we examine symptom severity from patient self-reports. For the retrospective cohort, we examine electronic health records (EHRs) for the existence of 28 potential adverse events during the 28-day period before and after each vaccination.

**Findings:** In the prospective cohort, 1,905 participants received COVID-19 vaccination; 899 received influenza vaccination. Focusing on those who received both vaccines yielded a total of 689 participants in the prospective cohort and 31,297 members in the retrospective cohort. *Questionnaire analysis*: For the COVID-19 vaccine, 39·7% [95% CI 36·4%–42·9%] of individuals reported no systemic reaction vs. 66·9% [95% CI 63·4%–70·3%] for the influenza vaccine. Individuals reporting a more severe reaction after influenza vaccination tended to likewise report a more severe reaction after COVID-19 vaccination (r=0·185, p<0·001). *Smartwatch analysis*: A statistically significant increase in heart rate and stress measure occurred during the first 3 days after COVID-19 vaccination, peaking 22 hours after vaccination with a mean increase of 4·48 (95% CI 3·94–5·01) beats per minute and 9·34 (95% CI 8·31–10·37) units in the stress measure compared to baseline. For influenza vaccination, we observed no changes in heart rate or stress measures. In paired analysis, the increase in both heart rate and stress measure for each participant was higher (p-value < 0·001) for COVID-19 vaccination than for influenza vaccination in the first 2 days after vaccination. On the second day after vaccination, participants had 1·5 (95% CI 0·68–2·20) more heartbeats per minute and 3·8 (95% CI 2·27–5·22) units higher stress measure, compared to their baseline. These differences disappeared by the third day after vaccination. *EHR analysis*: We found no elevated risk of non-COVID-19 or - influenza hospitalization following either vaccine. COVID-19 vaccination was not associated with an increased risk of any of the adverse events examined. Influenza vaccination was associated with an increased risk of Bell’s palsy (1·3 [95% CI 0·3–2·6] additional events per 10,000 people).

**Interpretation:** The more pronounced side effects after COVID-19 vaccination compared to influenza vaccination may explain the greater concern regarding COVID-19 vaccines. Nevertheless, our findings support the safety profile of both vaccines, as the reported side effects and physiological reactions measured by the smartwatches faded shortly after inoculation, and no substantial increase in adverse events was detected in the retrospective cohort.

**Funding:** This work was supported by the European Research Council, project #949850, and a Koret Foundation gift for Smart Cities and Digital Living.

**RESEARCH IN CONTEXT:** 

**Evidence before this study:** The unprecedented global impact of COVID-19 led to the rapid development and deployment of vaccines against the virus, including vaccines using novel mRNA technology. Despite the promising effectiveness of mRNA vaccines in preventing severe outcomes of COVID-19, concerns have been raised regarding the safety profile of these new vaccines. These concerns led to a notable global public reluctance to become vaccinated. By contrast, the seasonal influenza vaccine has been administered for decades with a well-established safety record and a high level of public acceptance. We searched Google Scholar, PubMed, and preprint services (including medRxiv, bioRrxiv, and SSRN) for studies comparing the safety profile of the two vaccines between March 1, 2023 (our study’s launch) and May 30, 2023, with no language restrictions, using the terms “safety of” AND (“COVID-19” OR “SARS-CoV-2”) AND (“vaccine” OR “BNT162b2 (Pfizer–BioNTech) mRNA vaccine”) AND “compared to” AND (“Influenza” OR “seasonal influenza” OR “flu”) AND “vaccine”. We found a study that compared the safety profile of the mRNA COVID-19 vaccine among 18,755 recipients with 27,895 recipients of the seasonal influenza vaccine using the WHO international database. The authors found a different safety pattern between the two vaccines with more systematic reactions following inoculation of the COVID-19 vaccine. Additionally, COVID-19 vaccines were associated with a higher risk of cardiovascular adverse events, while the influenza vaccine was associated with a higher risk of neurological adverse events. The remaining studies identified in our search compared the simultaneous administration of both vaccines to the administration of only COVID-19 vaccines. None of the studies conducted a paired analysis that compared reactions post-influenza vaccination and post-COVID-19 vaccination for the same individual; none examined the extent of physiological reaction (in terms of heart rate and heart rate variability) following the administration of COVID-19 or seasonal influenza vaccines; and none examined a cohort of individuals with data from before and after vaccination episodes or presented a comprehensive analysis to address concerns regarding the existence of potential rare adverse events following vaccination.

**Added value of this study:** We studied a prospective cohort of 5,079 participants in Israel (the PerMed study) from October 31, 2020 to September 30, 2022 and a retrospective cohort of 250,000 members of Maccabi Healthcare Services from July 31, 2021 and March 1, 2023. We examined reactions to BNT162b2 (Pfizer-BioNTech) mRNA COVID-19 vaccination (third or fourth shot) and to influenza vaccination. We compared the extent of reactions at the individual level, among individuals who received both vaccines separately. While the self-reported data and the continuous physiological measures from smartwatches revealed a higher rate of reactions following COVID-19 vaccination, these reactions faded soon after inoculation. We found no increase in risk of rare adverse events for either vaccine. We found a weak, albeit significant, correlation in the severity of the symptoms for the two vaccines (r=0·185, p<0·001): individuals who reported a more severe reaction after influenza vaccination tended to likewise report a more severe reaction after COVID-19 vaccination. We found no elevated risk of non-COVID-19 or - influenza hospitalization following the administration of either vaccine. COVID-19 vaccination was not associated with increased risk of any of the adverse events examined. Influenza vaccination was associated with an increased risk of Bell’s palsy (1·3 [95% CI 0·3–2·6] additional events per 10,000 people).

**Implications of all the available evidence:** Our study demonstrates the importance of accounting for continuous and objective surveillance of vaccines in both the clinical trial phase and the post-marketing phase, as it can aid in evaluating the safety profile of clinical trials and reduce vaccine hesitancy. The more pronounced side effects after COVID-19 vaccination compared to influenza vaccination may explain the greater concern regarding COVID-19 vaccines. Nevertheless, our findings support the safety profile of both vaccines, as the reported side effects and physiological reactions measured by the smartwatches faded shortly after inoculation, and no substantial increase in adverse events was detected in the retrospective cohort.

## INTRODUCTION

The unprecedented global impact of COVID-19 led to the rapid development and deployment of vaccines against the virus, including vaccines using novel mRNA technology. However, despite the promising safety profile and effectiveness of mRNA vaccines in preventing severe outcomes of COVID-19, there has been a notable global public reluctance to be vaccinated.^1, 2^ For example, in the United States nearly 20% of the population has received no COVID-19 vaccine doses.^3^ A key reason for COVID-19 vaccine hesitancy is concerns about the safety of the vaccine.^4, 5^

One strategy to address such concerns is by comparing the side effects of the mRNA vaccines to the side effects of the seasonal influenza vaccine, which has been administered for decades with a solid safety record and a high level of public acceptance. Hundreds of millions of vaccines for seasonal influenza and COVID-19 have been administered to date^6, 7^ but information comparing the safety of these vaccines is limited.

Currently, information on vaccine side effects is primarily collected through self-reporting methods, but these methods may be subject to bias and underreporting. To address this issue and provide a more comprehensive assessment of vaccine safety, extensive, continuous, and detailed monitoring of physiological changes in vaccinated individuals is necessary. Wearable devices, such as smartwatches, offer a promising solution. They enable continuous, detailed monitoring of physiological changes in vaccinated individuals, which can help identify vaccine-associated adverse events more effectively than self-reporting alone.

Wearable sensors have been shown to detect subtle medical conditions, such as atrial fibrillation, based on irregular heartbeats.^8^ Several studies have shown that heart metrics, including heart rate, heart rate variability, and resting heart rate, can indicate COVID-19 infection in the pre-symptomatic stage and thus can be used for real-time detection.^9–11^ These heart metrics have been previously reported to correlate with subjective symptoms after the COVID-19 vaccine.^12, 13^ In the context of COVID-19 vaccination, a number of studies using wearables have observed short-term changes in heart metrics following vaccination, even when such changes were not apparent to patients when self-reporting effects of vaccination.^14–18^

In this study, our goal is to evaluate the safety of mRNA COVID-19 booster vaccines by comparing their side effects with those of seasonal influenza vaccines, using self-reported data, physiological measurements from Garmin Vivosmart 4 smartwatches, and information from electronic health records (EHRs). We examined the short-term effects (lasting up to 42 days) of the BNT162b2 (Pfizer-BioNTech) mRNA COVID-19 booster vaccine and contrasted it with the seasonal influenza vaccine, by analyzing data from a prospective cohort of 5,079 participants in an observational trial, as well as a retrospective cohort of 250,000 randomly selected members from Maccabi Healthcare Services, Israel’s second-largest healthcare provider (which serves about 25% of the population).

## METHODS

### Cohorts

#### Prospective cohort

We studied a cohort of 5,079 participants from a prospective observational trial of individuals in the PerMed study^14–17, 19^ who received a third or fourth BNT162b2 mRNA COVID-19 vaccine and a seasonal influenza vaccine between July 31, 2021 and September 30, 2022 (Figure S2A, appendix p 13; see study protocol, appendix pp. 2-12) Participants filled out a daily questionnaire about clinical symptoms and wore a smartwatch that measured, among other factors, heart rate and heart rate variability-based stress.

Upon enrollment in the study, we gathered information on the participants’ sex, age, and pre-existing clinical risk factors. These underlying medical conditions included diabetes, hypertension, heart disease, chronic pulmonary conditions, weakened immune system, cancer, kidney failure, and a body mass index (BMI) greater than 30 (BMI is determined by dividing a person’s weight in kilograms by the square of their height in meters).

We paired observations so that we could compare vaccine reactions in each individual. Among the 5,079 participants in the prospective cohort, a total of 4,334 COVID-19 vaccine doses and 2,639 influenza vaccine doses were administered. For both types of vaccines, we had hourly smartwatch data on heart rate and a Garmin-computed stress measure based on heart rate variability (heart rate: 1,877 COVID-19 vaccine doses, 901 influenza vaccine doses; stress measure: 1,845 COVID-19 vaccine doses, 878 influenza vaccine doses) (Table S2 appendix p14). From these observations we extracted paired vaccine observations; these are observations for individuals who received both the COVID-19 vaccine (first or second booster) and at least one influenza vaccine.

#### Retrospective cohort

For the retrospective cohort, we examined anonymized EHRs of 250,000 randomly selected members of Maccabi Healthcare Services (Figure S2B, appendix p 13; see study protocol, appendix pp. 2-12). Eligible members were individuals 18 years and older who received both a third or fourth BNT162b2 mRNA COVID-19 vaccine and a seasonal influenza vaccine between July 31, 2021 and March 1, 2023. Individuals who were not members of Maccabi Healthcare Services throughout the entire study period were excluded.

### Study Design

#### Prospective cohort

For the prospective cohort, participants were asked to complete a daily survey via the PerMed mobile application.^19^ This survey compiled self-reported clinical symptoms from participants, using a predefined list of reactions observed in the BNT162b2 mRNA COVID-19 vaccine trial ^20^, while also allowing participants to freely report any additional symptoms they experienced. The survey was crafted based on potential symptoms that may follow infections with infectious diseases and respiratory illnesses, drawing on the International Classification of Diseases, Ninth Revision, and Clinical Modification (ICD-9) codes related to influenza, influenza-like illnesses, acute respiratory infections, RSV (respiratory syncytial virus), group A streptococcus, and COVID-19.

Throughout the study, from the moment they were recruited until the end, participants wore a Garmin Vivosmart 4 smartwatch. The smartwatch data was used to estimate the effects of vaccination on physiological measures such as heart rate and stress levels. Stress levels, computed by Garmin, range from 1 to 100 and are classified into four categories: resting (1-25), low (26-50), medium (51-75), and high (76-100).^21^ A higher stress level correlates with lower heart rate variability.^22, 23^ Heart rate data (beats per minute) was collected in intervals of 15 seconds while stress measurements were documented every three minutes.

Further information regarding the recruitment procedure, choice of smartwatch data analyzed, data collection architecture, and PerMed dashboard is provided elsewhere.^16^

#### Retrospective cohort

For the retrospective cohort, we examined anonymized EHRs of the patients. These records are automatically collected from various clinics and medical facilities nationwide and updated monthly in Maccabi Healthcare Services databases. The data is coded, anonymized, viewed, stored, and processed within the research room of the Maccabi Healthcare Services. Maccabi Healthcare Services uses the ICD-9 classification with procedures coded using Current Procedural Terminology codes. We obtained demographic details for each patient, along with diagnoses linked to 28 possible adverse events, as specified by ICD-9 codes.

### Outcomes and Statistical Analysis

#### Prospective cohort

Though the majority of participants only filled out the survey once daily, if there were multiple entries by a single participant within a day, only the final entry was considered. Since questionnaires cannot be updated after submission, participants were instructed to submit a new response in case of filing errors; we assumed that the last entry provided a more accurate representation of the participant’s entire day.

We defined a “baseline period” for each participant using their data (self-reported questionnaire and smartwatch) 7 days prior to vaccination. A participant’s “baseline” refers to the last questionnaire they submitted during the baseline period and smartwatch data for the entire baseline period. If a symptom was reported following the vaccination and was not noted during the baseline period, we interpreted this to be a side effect of the vaccine. Participants who did not complete the questionnaire during the baseline period were omitted from our analysis because we could not determine whether their symptoms existed prior to vaccination. We compared the baseline period to 7 and 14 days after vaccination inclusive of vaccination day (“post-vaccination” period), for discrete and continuous metrics, respectively. Since the U.S. Centers for Disease Control and Prevention (CDC) advises that vaccine side effects typically disappear after a few days, our analysis concentrates on the first 72 hours post-vaccination.^24^

We included participants who (1) submitted the questionnaire at least once during the baseline period, (2) completed the questionnaire at least once within 72 hours after vaccination, (3) provided wearable device measurements during the same day-of-week and time-of-day during their post-vaccination and baseline periods, and (4) received both the seasonal influenza and COVID-19 vaccine.

We excluded participants who provided data for only one day (either during the baseline or post-vaccination periods), with an exception if the participant provided data for the same hours-of-day and days-of-week during the baseline and post-vaccination periods.

We differentiated participants based on their self-reported intensity of symptoms as recorded in a questionnaire during the 72 hours following each vaccination. Participants were grouped into “No Reaction,” “Mild Reaction,” or “Severe Reaction” categories, based on their most severe symptom reported in the 72 hours following each vaccination. Consequently, if a participant experienced a single severe symptom for a day, and mild symptoms for the remaining three days post-vaccination, they were designated as having a severe reaction. The severity categorization could vary for participants after each administered vaccine dose.

In alignment with the CDC^25^ and the Pfizer clinical trial^20^, we categorized symptoms as follows:

- Mild symptoms: abdominal pain, feeling hot, back or neck pain, feeling cold, muscle pain, weakness, headache, dizziness, vomiting, sore throat, diarrhea, cough, leg pain, ear pain, loss of taste and smell, swelling of the lymph nodes, fast heartbeat, and hypertension;
- Severe symptoms: chest pain, dyspnea (shortness of breath), fever, confusion, and chills.

For participants who reported feeling hot and recorded their temperature, we divided them into two categories: above 38·9° C (fever) or below 38·9° C (feeling hot); in cases where the participant did not record their temperature, we assumed it to be below 38·9° C.

From the questionnaire data, we computed the proportion and corresponding 95% confidence interval (CI) of participants who reported experiencing each side effect in the post-vaccination period. The 95% CI for each side effect was calculated using a binomial distribution Binom(*n,p*), where *p* = the proportion of participants reporting the symptom, and *n* = number of participants eligible to be included in the self-reported questionnaire analysis. In cases where participants received more than one dose of the COVID-19 or the seasonal influenza vaccine during the study period, and were eligible to be included for both inoculations, each self-reported event (i.e., specific symptoms or no symptoms) was considered as 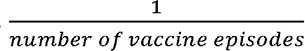 event to avoid over counting.

For the continuous Garmin smartwatch data, we compared measurements from the post-vaccination period to the corresponding day-of-week and hour-of-day measurement from the baseline period at the individual level. For example, we compare a participant’s Wednesday average 9 am heart rate with their previous Wednesday 9 am heart rate. If these data were not recorded (e.g., if a participant did not wear the smartwatch in the same period before and after vaccination), we excluded the participant from this analysis. Then, we aggregated each hour’s differences divided by all participants to calculate a mean difference and the associated 95% CI. We present this analysis starting a week before and after vaccination. In cases where participants received more than one dose of the COVID-19 or the seasonal influenza vaccines during the study period, and were eligible to be included for both inoculations, we calculated the average differences over all inoculations.

We also compared the difference of physiological changes between COVID-19 vaccination and seasonal influenza vaccination among individuals who received both vaccines. For each participant, we first calculated the daily mean changes in heart rate between the post-vaccination period and the baseline period. We did this separately for the mRNA COVID-19 booster vaccine and the seasonal influenza vaccine. Then we calculated the difference between these two mean values for each participant and each of the 7 days after inoculation. This is equivalent to a two-sided Welch’s t-test, which does not assume equal population variance.

#### Retrospective cohort

For each individual in the retrospective cohort who received both vaccines, we noted the existence in the EHR of 28 potential adverse events (Table S1, appendix pp 10-11) during the 28-day period before and after each vaccination. This set of adverse events was composed based on a previous large-scale study that examined the safety of the first and second (primary series) COVID-19 vaccine dose.^26^

We calculated the number of adverse events shown in patients’ EHRs before and after each vaccine. Consistent with a previous study^27^, we chose a time interval of 28 days to evaluate the potential short-term effects of each of the vaccines separately. Similar to large cohort studies,^16, 28^ we evaluated the risk differences using a self-as-control method that compared adverse events in the same patient in two periods: a baseline period of 28 days (from 35-8 days before vaccination) and a post-vaccination period of 28 days (0-27 days after vaccination). We used a 7-day buffer period between the baseline and post-vaccination periods in accordance with the guidelines of the Israeli Ministry of Health,^29^ which recommend that an individual should not receive inoculation if suffering from any apparent infection with severe symptoms, including a fever higher than 38°C. This 7-day buffer period was also consistent with the assumption made by a large-scale safety study in which non-vaccinated individuals were not eligible to serve as controls if the person experienced any health event in the week before the follow-up period.^26^

During outpatient visits in Maccabi’s clinics, past diagnoses also appear as part of the current visit’s diagnoses. Thus, in line with the previous study, for each adverse event we omitted individuals who were previously diagnosed with the same event in the year prior to the baseline period.^26^

We conducted a pairwise comparison for each individual, calculating the risk difference between pre- and post-vaccination values, which we denote *Y*^*i,j*^_*diff*_ for person *i* and event *j*. If an adverse event was recorded in the individual’s medical records in the post-vaccination period (i.e., 0-27 days post-vaccination) but not in the pre-vaccination period (35-8 days pre-vaccination), then the event is potentially associated with the vaccine or a random event, and we set *Y*^*i,j*^_*diff*_ = 1. If the converse was true and an adverse event appeared before vaccination but not after vaccination, the event is potentially a random event, and we set *Y*^*i,j*^_*diff*_ = -1. If a specific event was reported in both the pre- and post-vaccination periods, we assumed the event is not associated with the vaccine and we set *Y*^*i,j*^_*diff*_ = 0, and thus the individual was excluded from the analysis of event *j*. If an individual was found to be positive for COVID-19 during the post-vaccination period, we compared only the events recorded in the period between the inoculation and the recorded time of death and matched this period with the same time interval in the baseline period.

The risk difference for event *j* is the mean value of *Ȳ*^*j*^_*diff*_ calculated over all vaccinated individuals. This approach mirrors the standard estimation of risk differences in exposed and unexposed groups^30^, while taking into account the paired nature of the samples. To calculate the 95% CI for the difference without imposing any unknown distribution, we applied a non-parametric percentile bootstrap method with 10,000 repetitions, similar to previous safety studies^26, 28^ . In case an individual received more than one inoculation of COVID-19 vaccine or more than one inoculation of the seasonal influenza vaccine, for each repetition we chose randomly one of inoculations for each vaccine type.

### Ethical Approval

The prospective study was approved by MHS’ Helsinki institutional review board, protocol number 0122-20-MHS. All participants gave written informed consent to participate in the study and were advised both orally and in writing of the nature of the study. This study is part of a larger study (an observational clinical trial funded by a European Research Council grant) and is in accordance with the European Union General Data Protection Regulation: a cohort of 5,000 participants are recruited, download a dedicated mobile application, receive smartwatches, grant access to their medical records, and are followed for two years. Since the retrospective data was pseudonymized, the Helsinki institutional review board approved the use of this cohort data without requiring specific consent from Maccabi Healthcare Services members (protocol number 0122-20-MHS).

## RESULTS

### Prospective Cohort

#### Cohort characteristics

A total of 2115 participants received either the COVID-19 vaccine or the influenza vaccine during the study period (Table 1). Among these participants, 1905 received the COVID- 19 vaccine, while 899 received the influenza vaccine. In the paired sample of 689 participants receiving both vaccines, 355 (51·5%) were female, and age ranged from 20-85 years, with a median of 58. This is significantly higher than the median age in Israel, which is 30·5 years.^31^ Among the 689 participants, 357 (51·8%) reported that they had an underlying medical condition and 177 (25·7%) had a BMI of 30 or greater.

**Table 1.**
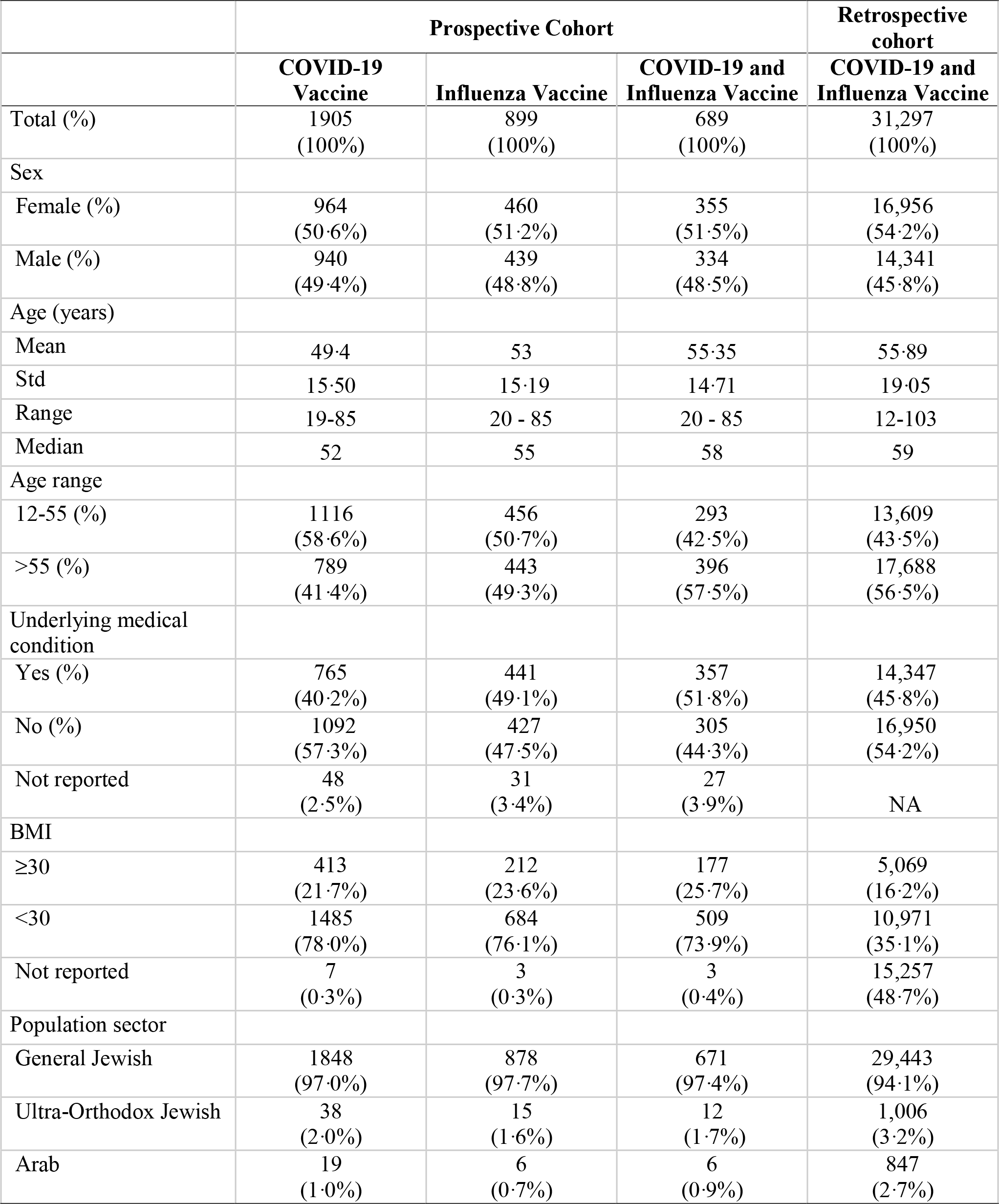
Description of cohort participants and self-reported reaction severity after COVID-19 booster vaccination and influenza vaccination for individuals receiving both vaccines

|  | **Prospective Cohort** |  |  | **Retrospective cohort** |
| --- | --- | --- | --- | --- |
|  | **COVID-19 Vaccine** | **Influenza Vaccine** | **COVID-19 and Influenza Vaccine** | **COVID-19 and Influenza Vaccine** |
| Total (%) | 1905<br>(100%) | 899<br>(100%) | 689<br>(100%) | 31,297<br>(100%) |
| Sex |  |  |  |  |
| Female (%) | 964<br>(50.6%) | 460<br>(51.2%) | 355<br>(51.5%) | 16,956<br>(54.2%) |
| Male (%) | 940<br>(49.4%) | 439<br>(48.8%) | 334<br>(48.5%) | 14,341<br>(45.8%) |
| Age (years) |  |  |  |  |
| Mean | 49.4 | 53 | 55.35 | 55.89 |
| Std | 15.50 | 15.19 | 14.71 | 19.05 |
| Range | 19-85 | 20 - 85 | 20 - 85 | 12-103 |
| Median | 52 | 55 | 58 | 59 |
| Age range |  |  |  |  |
| 12-55 (%) | 1116<br>(58.6%) | 456<br>(50.7%) | 293<br>(42.5%) | 13,609<br>(43.5%) |
| >55 (%) | 789<br>(41.4%) | 443<br>(49.3%) | 396<br>(57.5%) | 17,688<br>(56.5%) |
| Underlying medical condition |  |  |  |  |
| Yes (%) | 765<br>(40.2%) | 441<br>(49.1%) | 357<br>(51.8%) | 14,347<br>(45.8%) |
| No (%) | 1092<br>(57.3%) | 427<br>(47.5%) | 305<br>(44.3%) | 16,950<br>(54.2%) |
| Not reported | 48<br>(2.5%) | 31<br>(3.4%) | 27<br>(3.9%) | NA |
| BMI |  |  |  |  |
| ≥30 | 413<br>(21.7%) | 212<br>(23.6%) | 177<br>(25.7%) | 5,069<br>(16.2%) |
| <30 | 1485<br>(78.0%) | 684<br>(76.1%) | 509<br>(73.9%) | 10,971<br>(35.1%) |
| Not reported | 7<br>(0.3%) | 3<br>(0.3%) | 3<br>(0.4%) | 15,257<br>(48.7%) |
| Population sector |  |  |  |  |
| General Jewish | 1848<br>(97.0%) | 878<br>(97.7%) | 671<br>(97.4%) | 29,443<br>(94.1%) |
| Ultra-Orthodox Jewish | 38<br>(2.0%) | 15<br>(1.6%) | 12<br>(1.7%) | 1,006<br>(3.2%) |
| Arab | 19<br>(1.0%) | 6<br>(0.7%) | 6<br>(0.9%) | 847<br>(2.7%) |

When we paired observations for individuals who received both COVID-19 and influenza vaccines, we obtained 799 paired COVID-19 vaccine doses and 692 paired influenza vaccine doses for heart rate; and 779 paired COVID-19 vaccine doses and 672 paired influenza vaccine doses for the stress measure (Table S2, appendix p 14). These numbers are representative of 577 and 544 individuals with heart rate and stress data, respectively, who received both vaccines. Similarly, we had 881 paired daily self-reports on symptoms for 7 days pre- and post-COVID-19 vaccination and 727 pre- and post-influenza vaccination paired self-reports, corresponding to 621 individuals.

#### Questionnaire analysis

For the COVID-19 vaccine, 39·7% [95% CI 36·4%–42·9%] of individuals reported no systemic reaction; for the influenza vaccine, this fraction was 66·9% [95% CI 63·4%–70·3%] (Figure 1). The most common reactions reported for the COVID-19 vaccine were weakness (16·1% [95% CI 13·6%–18·5%] of vaccinated individuals), headache (13·1% [95% CI 10·9%–15·3%]), muscle pain (12·1% [95% CI 10·0%–14·3%]), fever (7·0% [95% CI 5·3%–8·7%]), and chills (5·7% [95% CI 4·2%–7·3%]). The most common reactions reported for the influenza vaccine were weakness (7·3% [95% CI 5·5%–9·2%]), headache (5·3% [95% CI 3·7%–7·0%]), muscle pain (3·6% [95% CI 2·3%–5·1%]), feeling cold (3·5% [95% CI 2·2%–5·0%]), and sore throat (3·0% [95% CI 1·8%–4·3%]).

**Figure 1.**
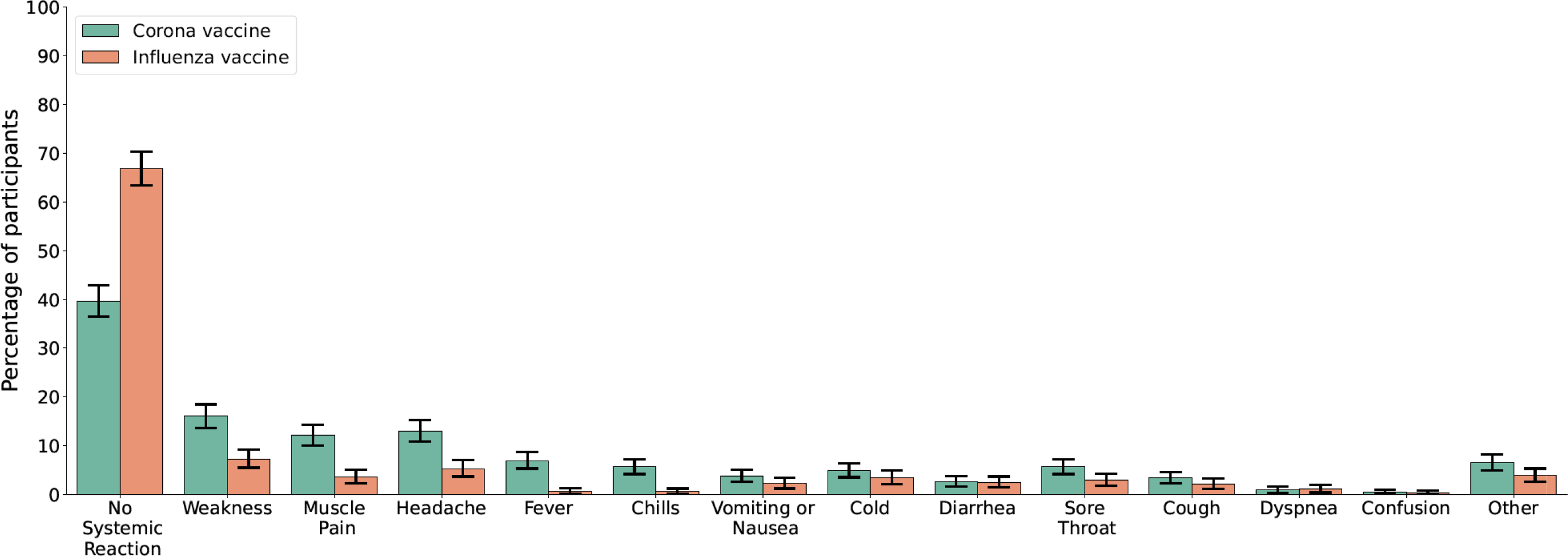
Reported symptoms following COVID-19 and influenza vaccination. Error bars represent 95% confidence intervals based on a binomial distribution.

We compared severity of symptoms for the 621 individuals who received both a COVID- 19 and influenza vaccine and who self-reported symptoms for both (Figure S3, appendix p 16). Using Spearman’s rank correlation test we found a weak, albeit significant, correlation in the severity of the symptoms for the two vaccines (r=0·185, p<0·001). Specifically, individuals who reported a more severe reaction after influenza vaccination tended to likewise report a more severe reaction after COVID-19 vaccination. Of the 621 individuals, 278 (44·8%) reported no reaction to either vaccine, 173 (27·8%) reported no reaction to the influenza vaccine and a mild reaction to the COVID-19 vaccine, and 36 (5·8%) reported no reaction to the influenza vaccine and a severe reaction to the COVID-19 vaccine. Similarly, 58 individuals (9·3%) reported a mild reaction to both vaccines, 38 individuals (6·1%) reported a mild reaction to the influenza vaccine and no reaction to the COVID-19 vaccine, and 25 individuals (4·0%) reported a mild reaction to the influenza vaccine and a severe reaction to the COVID-19 vaccine. Only 5 individuals (0·8%) reported a severe reaction to both vaccines. Of the 621 individuals, 321 (54·9%) reported the same level of symptom severity for both vaccines, and 259 (41·7%) reported a higher level of symptom severity for the COVID-19 vaccine than for the influenza vaccine. Only 46 individuals (7·4%) reported a lower level of symptom severity for the COVID-19 vaccine than for the influenza vaccine.

#### Wearables analysis

We examined mean pre- and post-vaccination differences in hourly heart rate and stress data for participants who received either the COVID-19 or influenza vaccine (Figure 2). Following the administration of the COVID-19 vaccine, we identified a statistically significant increase in heart rate during the first three days compared to the baseline period. This increase peaked 22 hours after vaccination, with a mean difference of 4·48 (95% CI 3·94–5·01) more beats per minute compared to baseline. By the sixth day post-inoculation, heart rate levels returned to baseline. A similar trend was observed for stress data: the increase in stress measure was statistically significant with a peak 22 hours after vaccination and a mean increase of 9·34 (95% CI 8·31–10·37) units in the stress measure. For influenza vaccination, no statistically significant changes in heart rate or the stress measure were observed.

**Figure 2.**
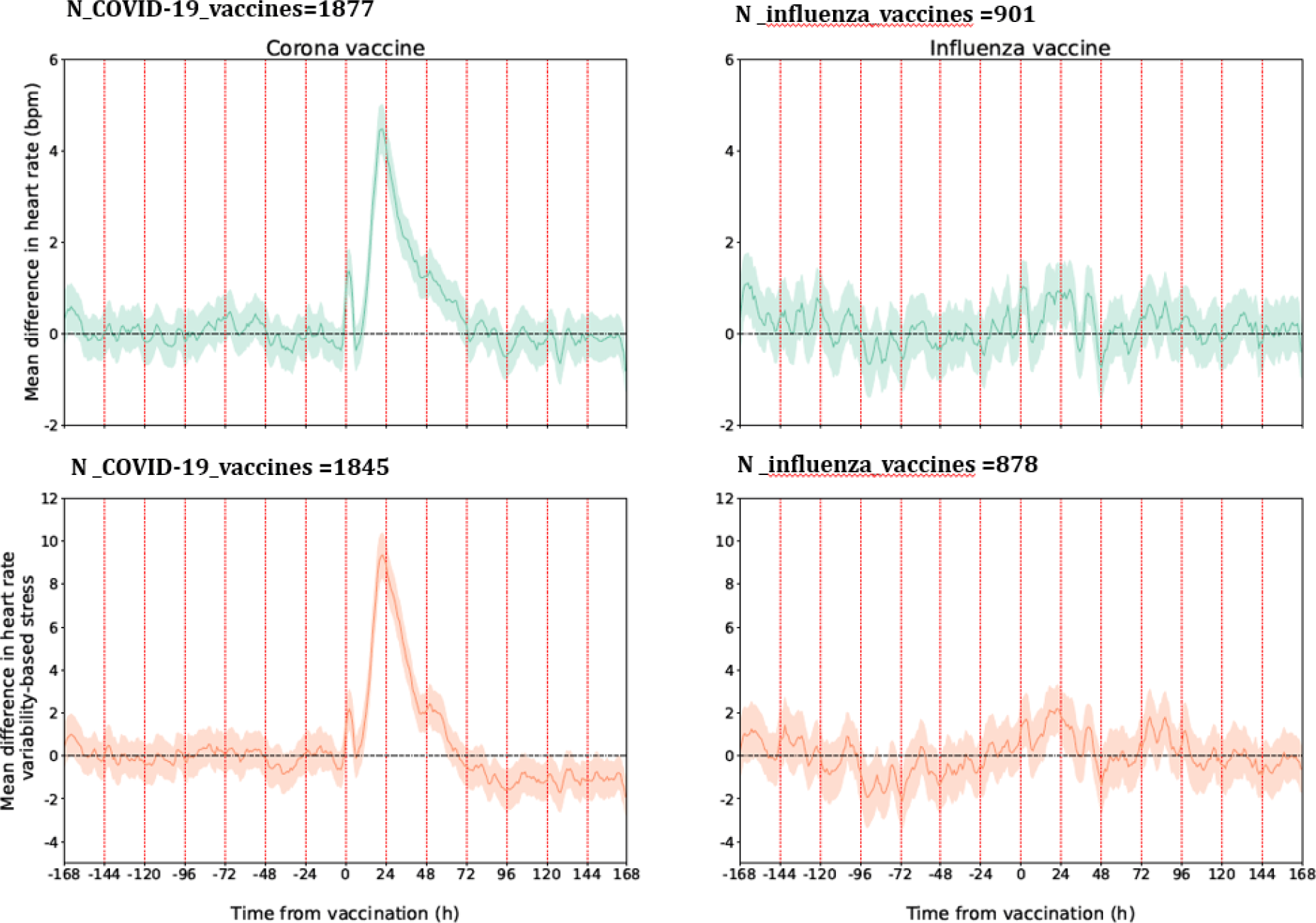
Mean difference in heart rate (in beats per minute) and stress measure (in points) between the post-vaccination and baseline periods after COVID-19 and influenza vaccinations. Shaded regions represent 95% confidence intervals.

To directly compare effects of the two vaccines we conducted a pairwise comparative analysis in which we examined daily mean changes in heart rate and the stress measure for individuals receiving both vaccines (Figure 3). For each such participant, we calculated the mean change in the indicator (either heart rate or stress) associated with COVID-19 vaccination compared to an individual’s baseline levels minus the mean change in the indicator associated with influenza vaccination compared to the individual’s baseline. Figure 3 shows that the increase in both heart rate and the stress measure for each participant was higher for COVID-19 vaccination than for influenza vaccination in the first two days after vaccination, and that this increase is statistically significant (p-value < 0·001). However, these differences were small: for example, in the second day after vaccination, mean heart rate was 1·5 (95% CI 0·68–2·20) beats per minute higher after COVID-19 vaccination than after influenza vaccination, compared to an individual’s baseline, and mean stress was 3·8 (95% CI 2·27–5·22) units higher. Moreover, these differences disappeared by the third day after vaccination: for the third through seventh days after vaccination there were no statistically significant differences in changes in these two indicators for the COVID-19 vaccine compared to the influenza vaccine.

**Figure 3.**
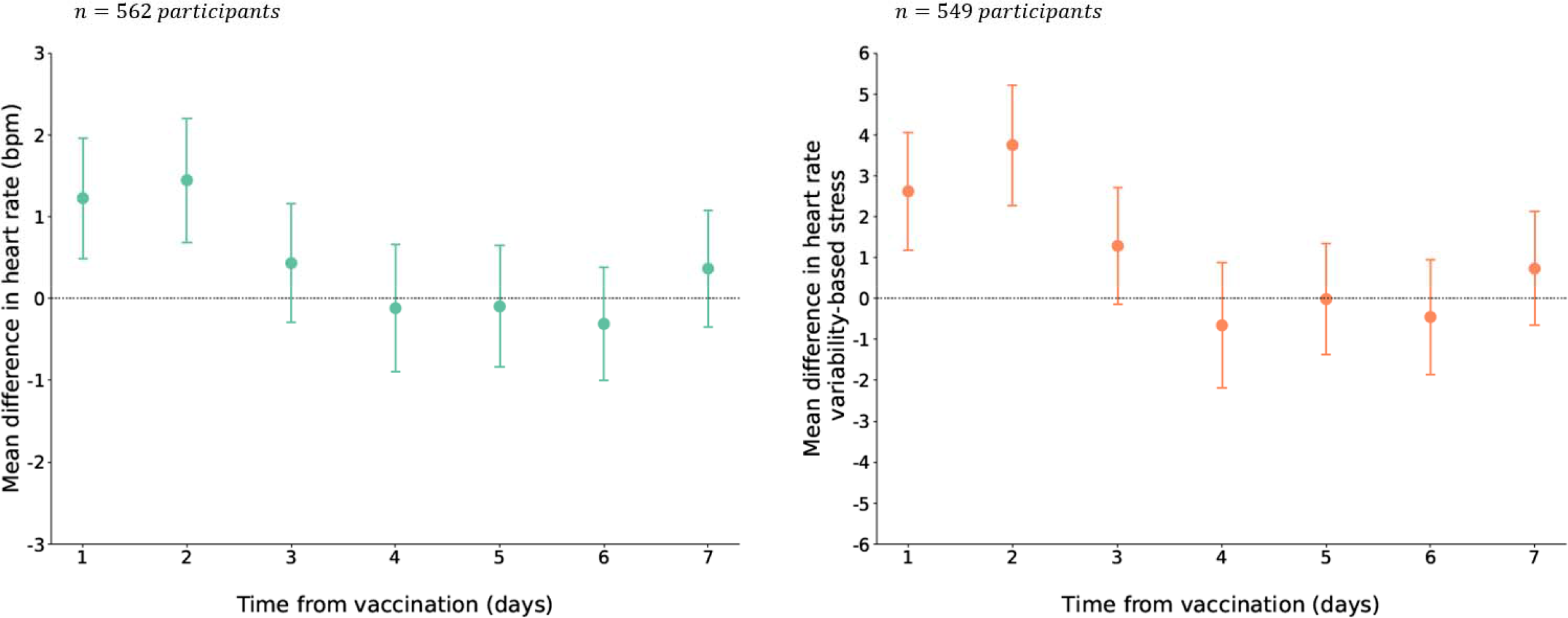
Paired analysis: daily mean changes in the smartwatch indicators for heart rate and the stress measure. For each participant, this was calculated as the mean change in the indicator (either heart rate or the stress measure) associated with COVID-19 vaccination ompared to an individual’s baseline minus the mean change in the indicator associated with influenza vaccination. Error bars represent 95% confidence intervals.

### Retrospective Cohort

#### Cohort characteristics

The retrospective cohort included 31,297 Maccabi members who received both vaccines during the study period. Of these individuals, 16,956 (54·2%) were female (Table 1). Age ranged from 12-103 years, with a median of 59. An underlying medical condition was listed in the medical record for 14,437 (45·8%) individuals, and 5,069 (16·2%) had a BMI of 30 or greater.

#### EHR analysis

We found no elevated risk of non-COVID-19 or -influenza hospitalizations following the administration of either vaccine (Table 2). Among the 31,297 individuals receiving both vaccines, 37 non-COVID-19 or -influenza hospitalizations occurred in the post-vaccination period compared to 51 in the baseline period for the COVID-19 vaccine and 32 non-COVID-19 or -influenza hospitalizations in the post-vaccination period compared to 30 in the baseline period for the influenza vaccine, corresponding to risk differences of -4·5 (95% CI: -10·2–1·3) and -0·6 (95% CI: -5·8–4·2) events per 10,000 vaccinated individuals, respectively.

**Table 2.** Adverse-events noted in the electronic health record during the 28-day period before and after each vaccination for the retrospective cohort

| Event | COVID-19 Vaccine |  |  |  | Influenza Vaccine |  |  |  |
| --- | --- | --- | --- | --- | --- | --- | --- | --- |
|  | Cohort<sup>*</sup> | Event before<sup>**</sup> | Event after<sup>**</sup> | Risk difference<sup>†</sup> | Cohort<sup>*</sup> | Event before<sup>**</sup> | Event after<sup>**</sup> | Risk difference<sup>†</sup> |
|  | *No. of persons* | *No. of events* | *No. of events* | *No. of events/10,000 persons* | *No. of persons* | *No. of events* | *No. of events* | *No. of events/10,000 persons* |
| **Non-COVID-19 or -influenza hospitalizations** | 31,286 | 51 | 37 | -4.5 (-10.2 – 1.3) | 31,287 | 32 | 30 | -0.6 (-5.8 – 4.2) |
| **Adverse events** |  |  |  |  |  |  |  |  |
| Acute kidney injury | 31,212 | 7 | 7 | 0.0 (-2.2 – 2.2) | 31,193 | 13 | 8 | -1.6 (-4.5 – 1.3) |
| Anemia | 29,636 | 165 | 118 | -15.9 (-27.0 – -4.7) | 29,230 | 151 | 155 | 1.4 (-10.3 – 13.0) |
| Appendicitis | 31,261 | 5 | 3 | -0.6 (-2.6 – 1.0) | 31,257 | 8 | 4 | -1.3 (-3.5 – 1.0) |
| Arrhythmia | 30,189 | 91 | 69 | -7.3 (-15.6 – 1.0) | 30,037 | 73 | 76 | -1.0 (-6.7 – 9.0) |
| Arthritis or arthropathy | 31,060 | 20 | 9 | -3.5 (-7.1 – 0.0) | 31,038 | 19 | 24 | 1.6 (-2.6 – 5.8) |
| Bell's palsy | 31,229 | 6 | 3 | -1.0 (-2.9 – 1.0) | 31,220 | 0 | 4 | 1.3 (0.3 – 2.6) |
| Cerebral venous sinus thrombosis | 31,287 | 0 | 1 | 0.3 (0.0 – 1.0) | 31,286 | 0 | 0 | .. |
| Deep vein thrombosis | 31,158 | 8 | 7 | -0.3 (-2.9 – 2.2) | 31,151 | 14 | 16 | 0.6 (-2.9 – 4.2) |
| Guillain-Barré syndrome | 31,283 | 0 | 0 | .. | 31,283 | 0 | 1 | 0.3 (0.0 – 1.0) |
| Heart failure | 31,004 | 37 | 19 | -5.8 (-10.6 – 1.3) | 30,939 | 20 | 22 | 0.6 (-3.6 – 4.8) |
| Herpes simplex virus infection | 31,131 | 22 | 15 | -2.2 (-6.1 – 1.6) | 31,109 | 22 | 16 | -1.9 (-5.8 – 1.9) |
| Herpes zoster virus infection | 31,046 | 31 | 18 | -4.2 (-8.7 – 0.0) | 30,965 | 35 | 17 | -5.8 (-10.3 – -1.3) |
| Intracranial hemorrhage | 31,253 | 3 | 3 | -0.0 (-1.6 – 1.6) | 31,243 | 4 | 4 | 0.0 (-1.9 – 1.6) |
| Ischemic stroke | 31,009 | 31 | 15 | -5.2 (-9.4 – 1.0) | 30,955 | 10 | 18 | 2.6 (-0.6 – 5.8) |
|  | Cohort<sup>*</sup> | Event before<sup>**</sup> | Event after<sup>**</sup> | Risk difference<sup>†</sup> | Cohort<sup>*</sup> | Event before<sup>**</sup> | Event after<sup>**</sup> | Risk difference<sup>†</sup> |
|  | *No. of persons* | *No. of events* | *No. of events* | *No. of events/ 10,000 persons* | *No. of persons* | *No. of events* | *No. of events* | *No. of events/ 10,000 persons* |
| Lymphadenopathy | 31,034 | 25 | 39 | 4.5 (-0.6 – 9.7) | 31,004 | 36 | 24 | -3.9 (-8.7 – 1.0) |
| Lymphopenia | 31,287 | 0 | 0 | .. | 31,287 | 0 | 0 | .. |
| Myocardial infarction | 31,169 | 15 | 13 | -0.6 (-3.8 – 2.6) | 31,141 | 9 | 14 | 1.6 (-1.3 – 4.8) |
| Myocarditis | 31,282 | 1 | 1 | -0.0 (-1.0 – 1.0) | 31,278 | 1 | 0 | -0.3 (1.0 – 0.0) |
| Neutropenia | 31,210 | 13 | 11 | -0.6 (-3.8 – 2.6) | 31,193 | 14 | 7 | -2.2 (-5.1 – 0.6) |
| Other thrombosis | 31,257 | 5 | 3 | -0.6 (-2.6 – 1.0) | 31,256 | 3 | 5 | 0.6 (-1.0 – 2.6) |
| Paresthesia | 30,752 | 77 | 51 | -8.5 (-15.6 – -1.3) | 30,639 | 68 | 67 | -0.3 (-7.8 – 7.2) |
| Pericarditis | 31,264 | 1 | 4 | 1.0 (-0.3 – 2.6) | 31,257 | 2 | 1 | -0.3 (-1.3 – 0.6) |
| Pulmonary embolus | 31,227 | 3 | 0 | -1.0 (-2.2 – 0.0) | 31,217 | 2 | 5 | 1.0 (-0.6 – 2.6) |
| Seizures | 31,262 | 5 | 5 | 0.0 (-1.9 – 1.9) | 31,244 | 1 | 2 | 0.3 (-0.6 – 1.6) |
| Syncope | 30,989 | 39 | 39 | 0.0 (-5.5 – 5.5) | 30,928 | 49 | 36 | -4.2 (-10.0 – 1.6) |
| Thrombocytopenia | 31,205 | 12 | 10 | -0.6 (-3.5 – 2.2) | 31,195 | 5 | 11 | 1.9 (-0.6 – 4.5) |
| Uveitis | 31,231 | 5 | 2 | -1.0 (-2.6 – 0.6) | 31,224 | 5 | 4 | -0.3 (-2.2 – 1.6) |
| Vertigo | 29,874 | 182 | 145 | -12.4 (-24.4 – -0.7) | 29,540 | 158 | 144 | -4.7 (-16.2 – 6.8) |

\* Individuals without previous diagnosis of the event before vaccination.
\*\* 28-day period before vaccination and after vaccination.
† The risk difference and confidence interval were estimated for each individual in a paired fashion with the use of a percentile bootstrap method with 10,000 repetitions.

We found no statistically significant increase in risk of the examined adverse events for COVID-19 vaccination (Table 2). Influenza vaccination was associated with an increased risk of Bell’s palsy (1·3 [95% CI 0·3–2·6] additional events per 10,000 people), but was not associated with any other increase in risk of the examined adverse events.

## DISCUSSION

Our analyses of data from patient questionnaires and smartwatches (in a prospective study) and from EHRs (in a retrospective study) comparing side effects of COVID-19 booster vaccination and seasonal influenza vaccination further support the safety of the first and second BNT162b2 booster vaccinations in eligible populations. In the patient questionnaires, more individuals reported mild or severe effects after the COVID-19 vaccine than after the influenza vaccine (60·3% vs. 33·1%). We found a weak but significant positive correlation in the severity of the symptoms for the two vaccines. From the smartwatch data we identified a small but statistically significant increase in heart rate and the stress measure in the three days after COVID-19 vaccination, with levels returning to normal within six days after vaccination. For influenza vaccination, no statistically significant changes in these measures were observed. Although our paired analysis of reactions to COVID-19 and influenza vaccination suggested that the differences are significant, they were small and do not indicate lack of safety. Importantly, for the retrospective cohort, our analysis of EHRs – which are based on diagnoses of physicians in clinics and hospitals –identified no significant change in risk of adverse events after either vaccine, except for an elevated risk of Bell’s palsy after the influenza vaccine, a rare event. This finding is in line with a recent large-scale study that also found this positive association.^32^ Despite this finding, the benefits of influenza vaccination in preventing severe outcomes far exceed this risk.

At both the aggregate level where we compared average changes in smartwatch measures pre- and post-vaccination for COVID-19 versus influenza vaccination, and at the individual level where we compared each patient’s changes in these measures, we found an increase in heart rate: 4·48 (95% CI 3·94–5·01) beats per minute higher at the aggregate level, and 1·5 (95% CI 0·68– 2·20) beats per minute higher 22 hours after vaccination when we performed a pairwise comparison. Previous studies have found that even small long-lasting increases in heart rate are associated with an increased risk of death.^33, 34^ However, we found that the increases in heart rate disappeared within six days after vaccination. We also found increases in the heart rate variability-based stress measure. Some details are available on how the Garmin stress measure is calculated,^35, 36^ but the exact algorithm is proprietary and is not fully disclosed. Although the stress measures returned to baseline within six days after vaccination, the clinical significance of the short-term increases in the stress measure is unknown.

Our analysis has several limitations. First, we considered only BNT162b2 (Pfizer-BioNTech) mRNA vaccines. Second, median cohort age was greater than the median age in Israel, reflecting the fact that older individuals are more likely to receive both influenza vaccination and a COVID-19 booster shot than younger individuals. It is possible that younger individuals might have a different side effects profile than found in our cohorts. Third, the Garmin smartwatches are not medical-grade wearable devices, although previous studies have demonstrated the accuracy of smartwatches in measuring heart rates^37, 38^ and our analysis considers relative heart rate values pre- and post-vaccination rather than absolute heart rate values.

Our analysis demonstrates the power of examining patient data from multiple sources. Although more individuals experienced side effects after COVID-19 vaccination than after influenza vaccination, as reflected both in patient self-reports and in smartwatch data, the differences in examined side effects detected by the smartwatches were small, the differences disappeared by three days after vaccination, and heart measures returned to their pre-vaccination baseline within six days after vaccination. Moreover, no statistically significant increase in the risk of adverse events, as reflected in EHRs, was associated with COVID-19 vaccination. Taken together, these findings support the safety of the first and second BNT162b2 (Pfizer-BioNTech) mRNA COVID-19 booster shots.

## Data Availability

According to this study MHS Helsinki and data utilization committees' guidelines, no patient-level data is to be shared outside the permitted researchers. Statistical analysis code will be available upon publication.

## Acknowledgments

This work was supported by the European Research Council, project #949850, and a Koret Foundation gift for Smart Cities and Digital Living.

## Authors’ contributions

Conception and design: DY, MY, ES and MLB. Collection and assembly of data: DY, MY, YL, and ES had access to the raw data and were responsible for verifying the data. Analysis and interpretation of the data: MY, DY, ES, YL, GQ, and MLB. Statistical analysis: MY, DY, ES and MLB. Drafting the article: MY, DY, ES, GQ and MLB. Critical revision of the article for important intellectual content: MY, DY, ES, GQ, ND and MLB. Final approval of the article: All authors. Obtaining funding: DY, ES, and MLB.

## Data sharing statements

According to this study’s MHS’s Helsinki and data utilization committees’ guidelines, no patient-level data is to be shared outside the permitted researchers. Statistical analysis code will be available upon publication.

## Supplementary Materials

### Appendix A – Study protocol Prospective part

#### Study Design

In this study we will analyze data that were already collected and will be collected as part of the PerMed study ^1^. Participants in the PerMed study are recruited for a period of two years, during which they are equipped with a Garmin Vivosmart 4 smartwatches and are asked to wear them as much as they could. In addition, participants install two applications on their mobile phones: an application that passively collects data from the smartwatch and a dedicated mobile application which allows participants to fill a daily questionnaire and to report their vaccine date and specific hour. In this study, we will consider for each participant, the 7-days period prior to any vaccination dose as the baseline period.

#### Participants

The inclusion criteria for the PerMed study includes those aged > 18 years. Individuals who are not eligible to give and sign a consent form of their free are excluded. In this study, we will analyze the data of participants aged 18 years and above, who reported receiving at least one dose of the BNT162b2 mRNA COVID-19 booster vaccine or seasonal influenza vaccine after joining the PerMed study. To recruit participants and ensure they complete all the study’s requirements, we will hire a professional survey company. Potential participants will be recruited through advertisements in social media, online banners, and word-of-mouth. The survey company is responsible for guaranteeing the participants meet the study’s requirements, in particular, that the questionnaires are filled daily, ensuring the smartwatches are charged constantly and worn properly, and assisting participants resolve technical problems.

#### Study procedures

Before participation in the study, all participants will be advised orally and in writing about the nature of the experiments and give written, informed consent. At this time, participants will be asked to complete an enrollment questionnaire that includes demographic information and health status. In addition, participants will be asked to install two applications on their mobile phones: an application that passively collects data from the smartwatch and the PerMed application, which allows participants to fill in the daily questionnaires. Participants will be given instructions regarding the self-reported symptoms questionnaires and how to operate the smartwatch, which they will wear as much as they can.

#### Enrollment questionnaire

All participants will fill a one-time enrollment questionnaire that includes demographic questions and questions about the participant’s health condition in general. Specifically, the questionnaire will include the following: age, gender, height, weight and underlying medical conditions (Listed in Table 1, main text). Other questions such as name, address, phone and email will be recorded and used by the survey company to contact the participants. The answers will be filled-in directly by the survey company to the study’s secured dashboard.

#### Monitoring device

Participants will be equipped with Garmin Vivosmart 4 smart fitness trackers. Among other features, the smartwatch provides all-day heart rate and heart rate variability and during-night blood oxygen saturation level tracking capabilities ^2^.

The optical wrist heart rate (HR) monitor of the smartwatch is designed to continuously monitor a user’s heart rate. The frequency at which heart rate is measured varies and may depend on the level of activity of the user: when the user starts an activity, the optical HR monitor’s measurement frequency increases.

Since heart rate variability (HRV) is not easily accessible through Garmin’s application programming interface (API), we use Garmin’s stress level instead, which is calculated based on HRV. Specifically, the device uses heart rate data to determine the interval between each heartbeat. The variable length of time between each heartbeat is regulated by the body’s autonomic nervous system. Less variability between beats correlates with higher stress levels, whereas an increase in variability indicates less stress ^3^. A similar relationship between HRV and stress was also seen in ^4, 5^.

The Pulse Ox monitor of the smartwatch uses a combination of red and infrared lights with sensors on the back of the device to estimate the percentage of oxygenated blood (peripheral oxygen saturation, SpO2%). The Pulse Ox monitor is activated each day at a fixed time for a period of four hours (the default is 2AM-6AM).

Examining the data collected in our study, we identified an HR sample roughly every 15 seconds, an HRV sample every 180 seconds, and an SpO2 sample every 60 seconds.

While the Garmin smartwatch provides state-of-the-art wrist monitoring, it is not a medical-grade device, and some readings may be inaccurate under certain circumstances, depending on factors such as the fit of the device and the type and intensity of the activity undertaken by a participant ^6–8^.

#### Vaccination questionnaire

The vaccination questionnaire we will use includes the following question:

COVID-19 vaccination – date, time and dose number. [note, this is for validation as vaccination data are reported in the EMR]

#### Daily questionnaires

All participants will complete the daily self-reported questionnaire in a dedicated application (the PerMed mobile application). The daily questionnaire we will use includes the following questions:

How is your mood today? • Awful (-2)• Bad (-1)• OK (0)• Good (1)• Excellent (2)

How would you describe the level of your stress during the last day?• Very Low (-2)• Low (-1)• Medium (0)• High (1)• Very high (2)

How would you define your last night sleep quality?• Awful (-2)• Bad (-1)• OK (0)• Good (1)• Excellent (2)

Try to remember how many minutes of sports activity you performed on the last day?

Have you experienced one or more of the following symptoms in the last 24 hours?• My general feeling is good, and I have no symptoms• Heat measured above 37·5• Cough• Sore throat• Runny nose• Headache• Shortness of breath• Muscle aches• Weakness / fatigue• Diarrhea• Nausea / vomiting• Chills• Confusion• Loss of sense of taste / smell• Another symptom.

#### Data Storage

Data collected from the mobile phone application and from the smartwatches will be stored on a secure server within Tel Aviv University facilities. The server runs a CentOS operating system and is located in Software Engineering Building at Tel Aviv University. This server is protected behind the university’s firewall and is not connected to external networks. In addition, a secure connection through an SSL protocol and a trusted certificate will be obtained for the transfer of information from the mobile phone application into the secured server.

Access will be restricted to investigators in the study. The information from the mobile application will be stored in a structured manner on the secured server without any explicitly identifying information (name, ID number, email). Each participant will be assigned a coded participant number that will be used to identify the subject in the database. The code with the identified information will be stored in an encrypted form on a separate secured server that only the research manager will have access to. Access to all servers is restricted with username and password.

All (non-digital) questionnaires and signed informed consent documents will be stored in a secured cabinet in Tel Aviv University, to which only the research manager and the principal investigators will have access. No data collected as part of the study will be added to individuals’ medical charts.

#### Data processing

We will perform several preprocessing steps. Concerning the daily questionnaires, in cases where participants will fill in the daily questionnaire more than once on a given day, only the last entry for that day will be considered, as it is reasoned that the last one likely best represented the entire day. Self-reported symptoms that are entered as the free text will be manually categorized. With regard to the smartwatch physiological indicators, data will first be aggregated per hour (by taking the mean value). Then, to impute missing values, we will perform a linear interpolation. Finally, data will be smoothed by calculating the moving average value using a five-hour sliding window.

#### Data Analysis and inclusion criteria

The questionnaire data will be preprocessed by manually categorizing any self-reported symptom entered as free text. If participants filled out the questionnaire more than once in one day, the last entry from that day was used in the analysis as it is likely more representative of the past day. Smartwatch data will be preprocessed as follows. First, we will compute the mean value of each hour of data. We will then perform a linear interpolation to impute missing hourly means. Lastly, we will smooth the data by calculating the five-hour moving average.

For each participant and each of the vaccines, we define the 7-day period prior to vaccination as the baseline period. For the analyses involving self-reported questionnaires, we will include participants who submitted at least one questionnaire during the baseline period and at least one questionnaire during the seven days post-vaccination. The two questionnaires are required to understand the appearance of new reactions following vaccination. For the analyses involving smartwatch indicators, we will include participants who had at least one overlapping period of data (i.e., same day of the week and same hour during the day) during their baseline and post vaccination periods. The overlapping periods are required for computing the change in indicator values between the baseline and post-vaccination periods.

To compare the changes in specific smartwatch indicators (heart rate, HRV-based stress, resting heart rate, and step counts) over the 0-42 days post vaccination, with those of the baseline period, we will perform the following steps. First, for each participant and each hour during the seven days post-vaccination, we will calculat the difference between that hour’s indicator value and that of the corresponding hour in the baseline period (keeping the same day of the week and same hour during the day). Then, we will aggregate each hour’s differences over all participants to calculate a mean difference and the associated 95% confidence interval, which is analogous to a one-sided t-test with a significance level of 0·05. To determine the statistical significance of daily differences between the baseline and post vaccination period, we will calculate the mean daily difference for each participant and then used a one-sample t-test for each day. To compare the difference of physiological changes between COVID-19 vaccination and seasonal influenza vaccination among individuals who received both vaccines. For each participant, we will first calculate the daily mean changes in heart rate between the post-vaccination period and the baseline period. We will do this separately for the mRNA COVID-19 booster vaccine and the seasonal influenza vaccine. Then we will calculate the difference between these two mean values for each participant and each of the 7 days after inoculation. This is equivalent to a two-sided Welch’s t-test, which does not assume equal population variance.

To understand the extent of new reactions, post vaccination, we will first note any pre-existing signs and symptoms reported in the last completed questionnaire during the baseline period. Next, we will calculate the percentage of participants who reported new (i.e., not pre-existing) systemic reactions in the 7-day period after vaccination from the following list: fatigue, headache, muscle pain, cold, fever, sore throat, cough, chills, vomiting or nausea, diarrhoea, dyspnoea, confusion, loss of taste and smell, Shortness of breath. Participants could also report any other symptoms using free text. For each reaction we use a binomial distribution to determine a 95% confidence interval.

#### Potential Risks & Risk management

No specific risks arising from the smartwatches are expected, as the device is already commercialized with no known adverse reactions. The main risk in this study is the leakage of private data which we intend to manage as we describe in the following section.

#### Privacy/Confidentiality

Results from this study will be handled at an aggregated level. Individual data records will remain confidential and will not be published or shared with any third party. Signed and dated informed consent forms, as well as data recording sheets (e.g., case report forms) will be stored in locked cabinets during the study and following its completion. A file containing the personal details of the participants will be coded to help preserve confidentiality and will be separated from all other data collected throughout the study. This file will be kept by the principal investigator. Data will be stored on computers in password-protected files.

The data obtained from the smartwatch used in this study will be linked to a coded participant number. The smartwatch does not include a GPS. The data collected by the PerMed application will arrive directly to PerMed back-end servers and will be stored securely.

### Retrospective part

#### Description of the data

Data will be extracted from the Maccabi Healthcare Services (MHS) database. MHS is a nationwide health plan (payer-provider) representing a quarter of the population in Israel. The MHS database contains longitudinal data on a stable population of 2·2 million people since 1993 (with <1%/year moving out). Data are automatically collected and includes comprehensive laboratory data from a single central lab, full pharmacy prescription and purchase data, and extensive demographic data on each patient. MHS uses the International Classification of Diseases, Ninth Revision, Clinical Modification (ICD-9-CM) coding systems as well as self-developed coding systems to provide more granular diagnostic information beyond the ICD codes. Medications are coded according to the Israeli coding system with translations to anatomical therapeutic chemical (ATC) classification system wherever available. Procedures are coded using Current Procedural Terminology (CPT) codes. We will access to the following data for each patient:

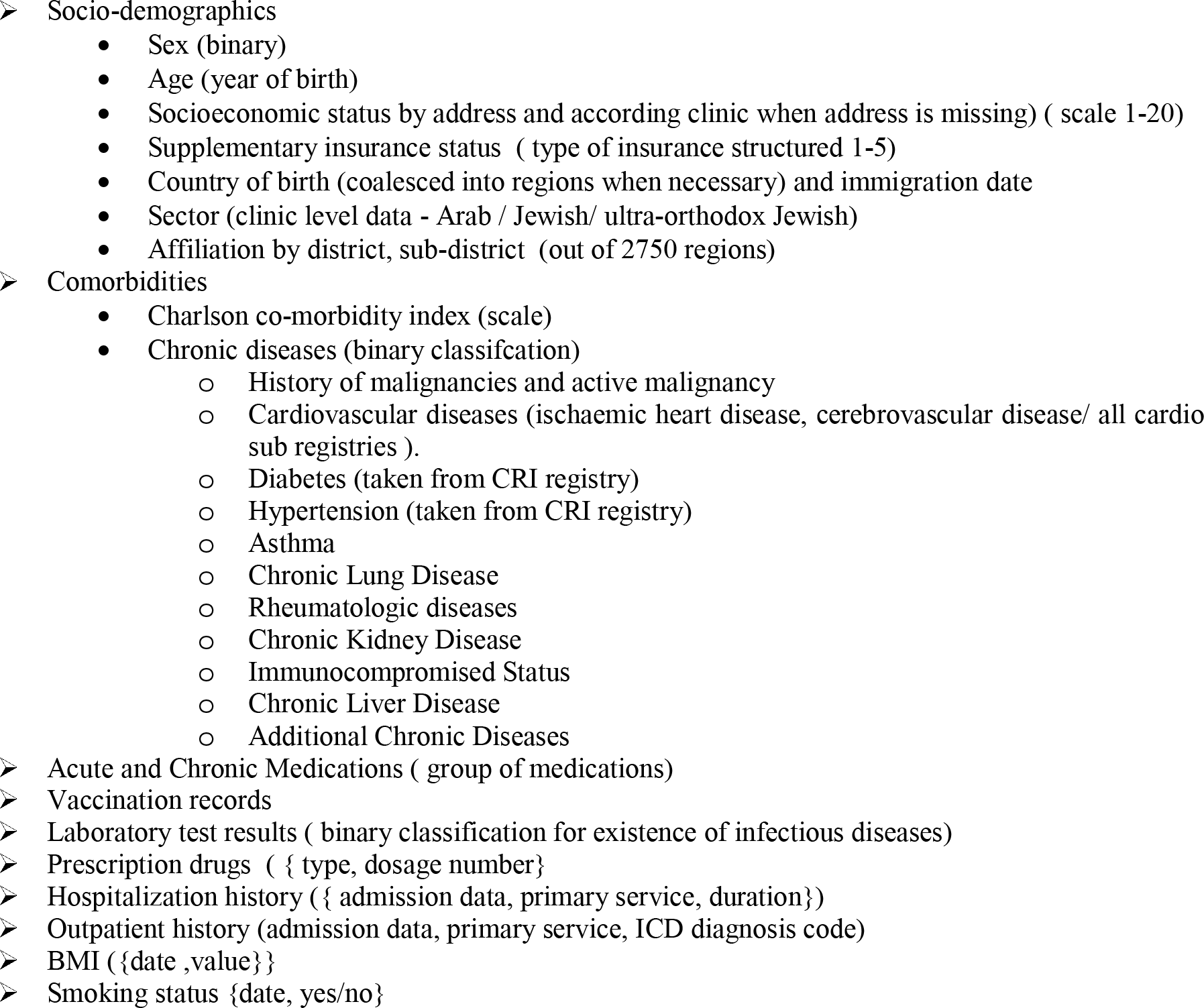

#### Data collection and storage

We will receive access to the data from the medical records of 250,000 random members of Maccabi and the 5,000 participants from the prospective cohort. MHS is a nationwide health plan (payer-provider) representing a quarter of the population in Israel. The MHS database contains longitudinal data on a stable population of 2·2 million people since 1993 (with <1%/year moving out). Data are automatically collected and includes comprehensive laboratory data from a single central lab, full pharmacy prescription and purchase data, and extensive demographic data on each patient. MHS uses the International Classification of Diseases, Ninth Revision, Clinical Modification (ICD-9- CM) coding systems as well as self-developed coding systems to provide more granular diagnostic information beyond the ICD codes. Medications are coded according to the Israeli coding system with translations to anatomical therapeutic chemical (ATC) classification system wherever available. Procedures are coded using Current Procedural Terminology (CPT) codes.

As for the medical data, we will receive access to the EMR data after the following pseudonymisation procedures:

1. Healthcare identification number of the members will be coded.
2. Only year of birth is provided
3. Free text is removed. This means any text that was types/recorded/scanned manually by healthcare staff, and is not structured in the electronic system. This includes any documented conversations between healthcare staff and patient or summary of from meetings.
4. No audio, photos including scanned text, or video contents are provided.
5. The address of the members is not detailed, and only the statistical area is provided (Israel is stratified into 2733 statistical areas with around 2500-5000 individuals per region).

The data access of the retrospective part will be conducted at the MHS. The data are coded, viewed, stored and process only within the Maccabi research room. The researchers will connect to the research room via MD Clone platform, which is approved by the Ministry of Health. The user connects through a secure connection using Lightweight Directory Access Protocol and two factor authentication system.

#### Potential adverse events

We will examine 28 potential adverse events (Table S1) that were previously investigated in the context of COVID- 19 vaccination ^9, 10^.

**Table S1.** ICD-9 codes of the examined potential adverse events in the retrospective cohort

| Event | ICD-9 Code |
| --- | --- |
| Acute kidney injury | 584.[5-9]* |
| Anemia | 28[0,1,3,4,5]* |
| Appendicitis | 54[0-2]*, 47* |
| Arrhythmia | 427*, 426* |
| Arthritis or arthropathy | 713*, 714.9*, 716.[4-9]*, 718.9, 719.[0,1,6,8,9]* |
| Bell's palsy | 351.0* |
| Cerebral venous sinus thrombosis | 325 |
| Deep vein thrombosis | 451, 451.[1-9]*, 453.[1,4]*, 453.8[0,2,3,4,5,6,7,8,9], 671.[3,4]* |
| Guillain-Barré syndrome | 357.0 |
| Heart failure | 428* |
| Herpes simplex virus infection | 054* |
| Herpes zoster virus infection | 053* |
| Intracranial hemorrhage | 43[0,1,2]* |
| Ischemic stroke | 433, 433.[0,1,2,3,8,9], 433.[0,1,2,3,8,9]1, 434*, 362.3[1-3], 436* |
| Lymphadenopathy | 785.6*, 683*, 289.[2,3]* |
| Lymphopenia | 288.5* |
| Myocardial infarction | 410* |
| Myocarditis | 422*, 429.0*, 398.0*, 391.2* |
| Neutropenia | 288.0, 288.0[0,3,4,9] |
| Other thrombosis | 444*, 557.[0,9]*, 557, 452*, 453, 453.[0,1,2,3,4,9]*, 453.[7,8], 453.[7,8][2-9], 437.6* |
| Paresthesia | 782.0* |
| Pericarditis | 420* |
| Pulmonary embolus | 415.1*, 673.[2,8]* |
| Seizures | 345.[2,3]*, 780.3, 780.39 |
| Syncope | 780.2*, 992.1* |
| Thrombocytopenia | 287.2*, 287.3, 287.3[0,1,3,9], 287.5 |
| Uveitis | 360.12, 362.18, 363.0*, 363.2[0,1,2], 363.1*, 364.[0,1,2,3]*, 053.22, 054.44, 091.5*, 098.41, 115.92 |
| Vertigo | 780.4*, 78.81, 386.1[0,1,9], 386.2, 438.85 |

\* Any of the possible ICD-9 combinations with a match

## Appendix B – Data collection platform and data access

### Architecture

The data collection platform contains several components that interact with each other (see Figure 2):

- **The PerMed application** – This application is installed on each participant’s phone to collect sensors data and the self-reported daily questionnaires. It also handles the smartwatch pairing. The current version of the application supports both Android and iOS devices.
- **The smartwatch** - send the data to the Garmin Connect app on the smartphone, which then sends these data to Garmin’s server.
- **The smartwatch application** – This application (currently Garmin) receives information from the smartwatch via Bluetooth and transmits it to the company’s server. In addition, it provides a convenient interface for displaying the participant’s smartwatch information.
- **The app server** – The webserver handles the database connectivity using REST API pages. It enables the server to authenticate users as they launch the application and write records to the database. A MySQL server stores the sensors’ raw data and the answers to the daily questionnaires. At last, there is a batch processes running on the server that sends app notifications (daily reminder to fill the questionnaire).
- **The dashboard server** - hosts the dashboard pages, which assist in monitoring the quality of the information and controlling the experiment. The dashboard has access to participant information and signals indicating whether questionnaires were completed and the smart watch was worn without seeing its content directly. A batch process is responsible for aggregating raw data for dashboard statistics.
- **The smartwatch server** - A MySQL server stores the smartwatch data. A batch process is responsible for collecting the data from the Garmin server.

**Figure S1.**
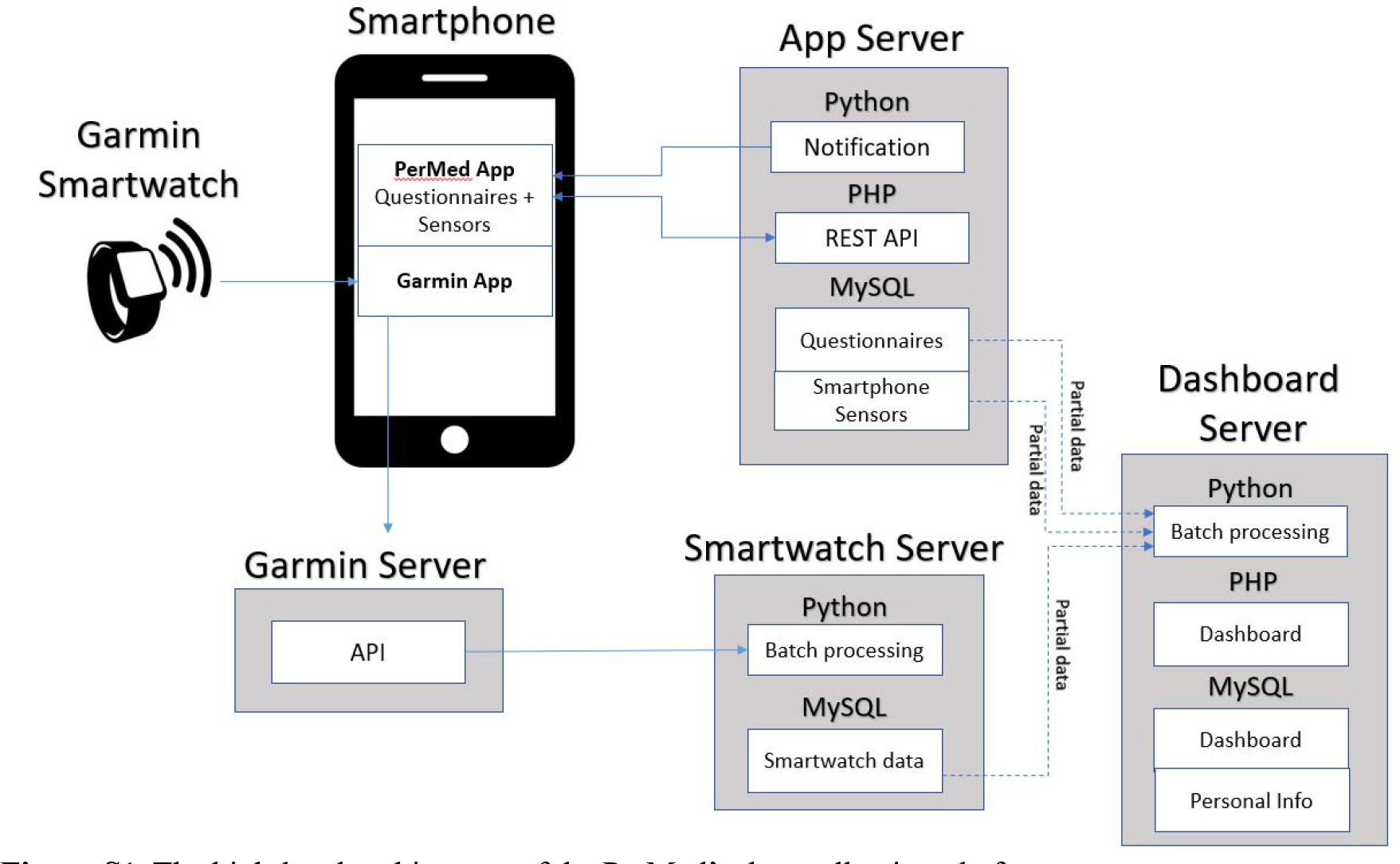
The high-level architecture of the PerMed’s data collection platform.

### The PerMed Dashboard

Participants will be recruited by a qualified external recruitment team headed by Tel Aviv University personal. The team receive limited information essential control the experiment. Thus, we developed a dedicated dashboard for monitoring the quality of the information and control the experiment. This dashboard aims to identify data collection issues such as participants who did not fill the daily questionnaires or participants who did not charge the battery of their smartwatches. The dashboard also helps us identify problems that were not related to participants’ cooperation, such as bugs in the mobileapp. This identification allow us to respond faster and provide timely solutions.

The Type of Data Collected and data access

Data collected by the platform arrive from four primary sources:

**Enrolment questionnaire -** data were collected from a one-time enrollment questionnaire that includes basic personal characteristics such as socio-demographic information (e.g., age, gender, height, weight), general habits, health status, and a short Big Five personality questionnaire.
**Daily questionnaire** – consists of questions on 1) wellbeing, 2) general health condition, 3) symptoms observed, 4) test results to diagnose infectious diseases, 4) vaccination or medicationconsumption (if relevant to the study question).
**Smartphone sensor data** – consist of location, Wi-Fi, Bluetooth, screen, and activity.
**Smartwatch data** - consist of heart rate data, accelerometer and gyroscope information and measures based on these data including active minutes, steps, distance, calories, and sleep level classification, including light, deep, REM, and awake periods.

The current research, aims to explore the safety of vaccination, is part of a larger study. Raw accelerometer data, mobile activity and GPS locations are generally considered sensitive information. In accordance with the data minimization principle, we did not extract these type of data for this vaccination safety research.

## Appendix C – Prospective study participants’ adherence

We employed a professional survey company to recruit participants and ensure they adhere to the study requirements. Participant recruitment was performed via advertisements on social media and word-of-mouth. Each participant signed an informed consent form after receiving a comprehensive explanation on the study. Then, participants completed a one-time enrollment questionnaire, were equipped with Garmin Vivosmart 4 smartwatches, and installed two applications on their mobile phones: (1) the PerMed application ^1, 11, 12^, which collects daily self-reported questionnaires, and (2) an application that passively records smartwatch data. Participants were asked to wear their smartwatches as much as possible. The survey company ensured that participants’ questionnaires were filled at least twice a week, that their smartwatches were charged and properly worn, and that any technical problems with the mobile applications or smartwatch were resolved. Participants were monitored through the mobile application and smartwatches for a period of at least 49 days, starting seven days before vaccination. Participants also granted full access to their EMR data.

We implemented several preventive measures to minimize participant attrition and discomfort as a means to improve the quality, continuity and reliability of the collected data. First, each day, participants who did not fill their daily questionnaire by 7 pm received a reminder notification through the PerMed application. Second, we developed a dedicated dashboard that allowed the survey company to identify participants who repeatedly neglected to complete the daily questionnaire or did not wear their smartwatch for extended periods of time; these participants were contacted by the survey company (either by text message or phone call) and encouraged to better adhere to the study protocol. Third, to strengthen participants’ engagement, a weekly personalized summary report was generated for each participant, which was available inside the PerMed application. Similarly, a monthly newsletter with recent findings from the study and useful tips regarding the smartwatch’s capabilities was sent to the participants. At the end of the study, participants will receive all personal insights that were obtained and can keep the smartwatch as a gift.

## Appendix D – Study design scheme

**Figure S2.**
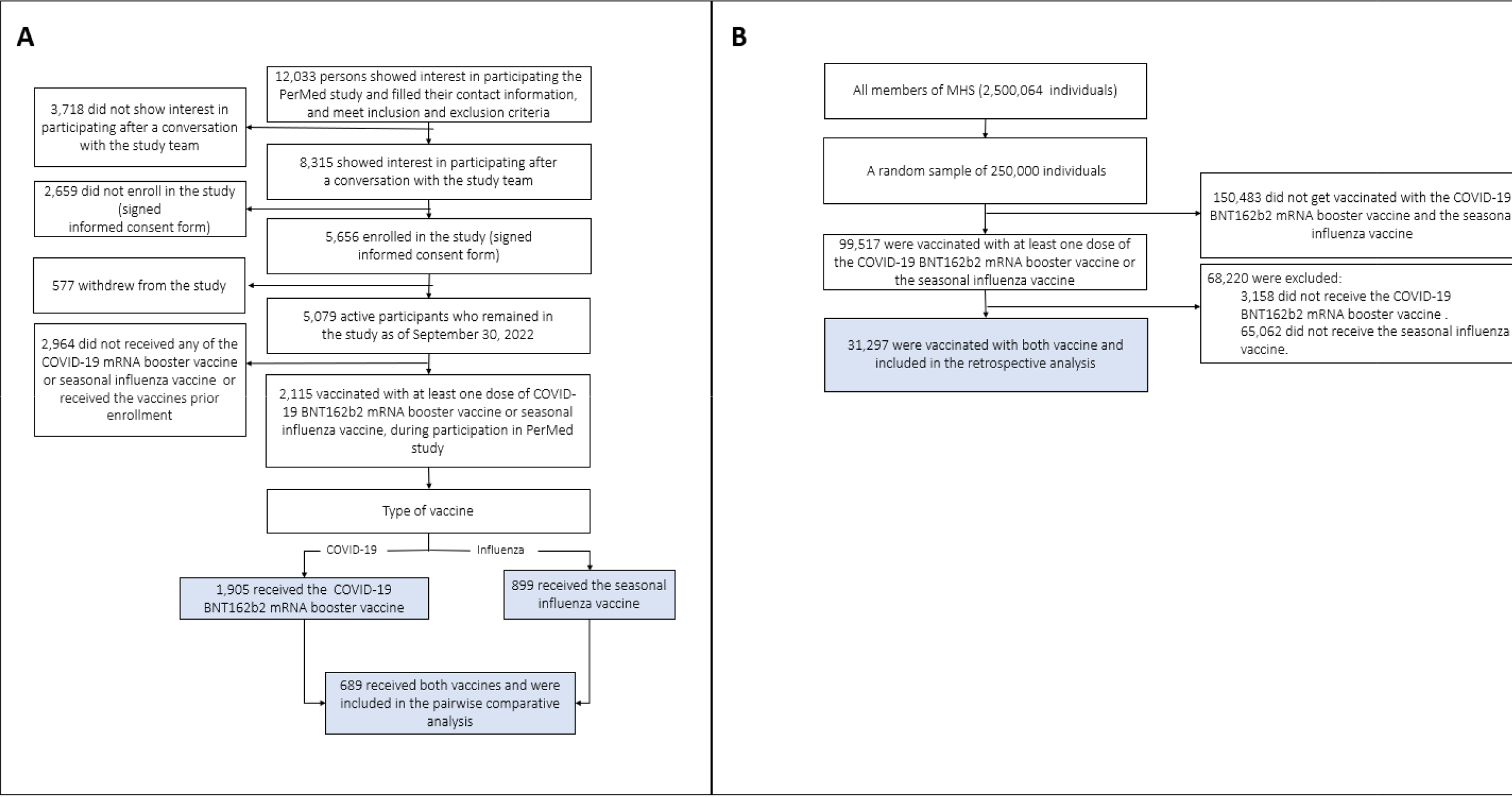
Trail profile. (A) prospective study, (B) retrospective study

**Table S2.** Number of prospective vaccine cohort observations *

| Vaccine type | No. vaccines | Hourly smartwatch data differences following vaccination |  |  |  | Daily smartwatch data differences following vaccination |  |  |  | Reported symptoms 7 days following vaccination |  |
| --- | --- | --- | --- | --- | --- | --- | --- | --- | --- | --- | --- |
|  |  | Heart Rate (Smoothed) |  | Heart Rate Variability (Smoothed) |  | Heart Rate (Smoothed) |  | Heart Rate Variability (Smoothed) |  |  |  |
|  |  | No. obser-<br>vations | No. paired<br>observations | No. obser-<br>vations | No. paired<br>observations | No. obser-<br>vations | No. paired<br>observations | No. obser-<br>vations | No. paired<br>observations | No. obser-<br>vations | No. paired<br>observations |
| COVID-19 (3<sup>rd</sup> and/or 4<sup>th</sup> dose) | 4334 | 1877 | 799 | 1845 | 779 | 1888 | 804 | 1858 | 784 | 2165 | 881 |
| Influenza | 2639 | 901 | 692 | 878 | 672 | 905 | 698 | 881 | 677 | 959 | 727 |
| Number of participants vaccinated with both vaccines |  | 577 |  | 544 |  | 562 |  | 549 |  | 621 |  |

\*Observations represent time series data following vaccination. Paired observations are those for which the individual received both a COVID-19 vaccination (3<sup>rd</sup> or 4<sup>th</sup> dose) and an influenza vaccination.

## Appendix E – Additional results

**Figure S1.**
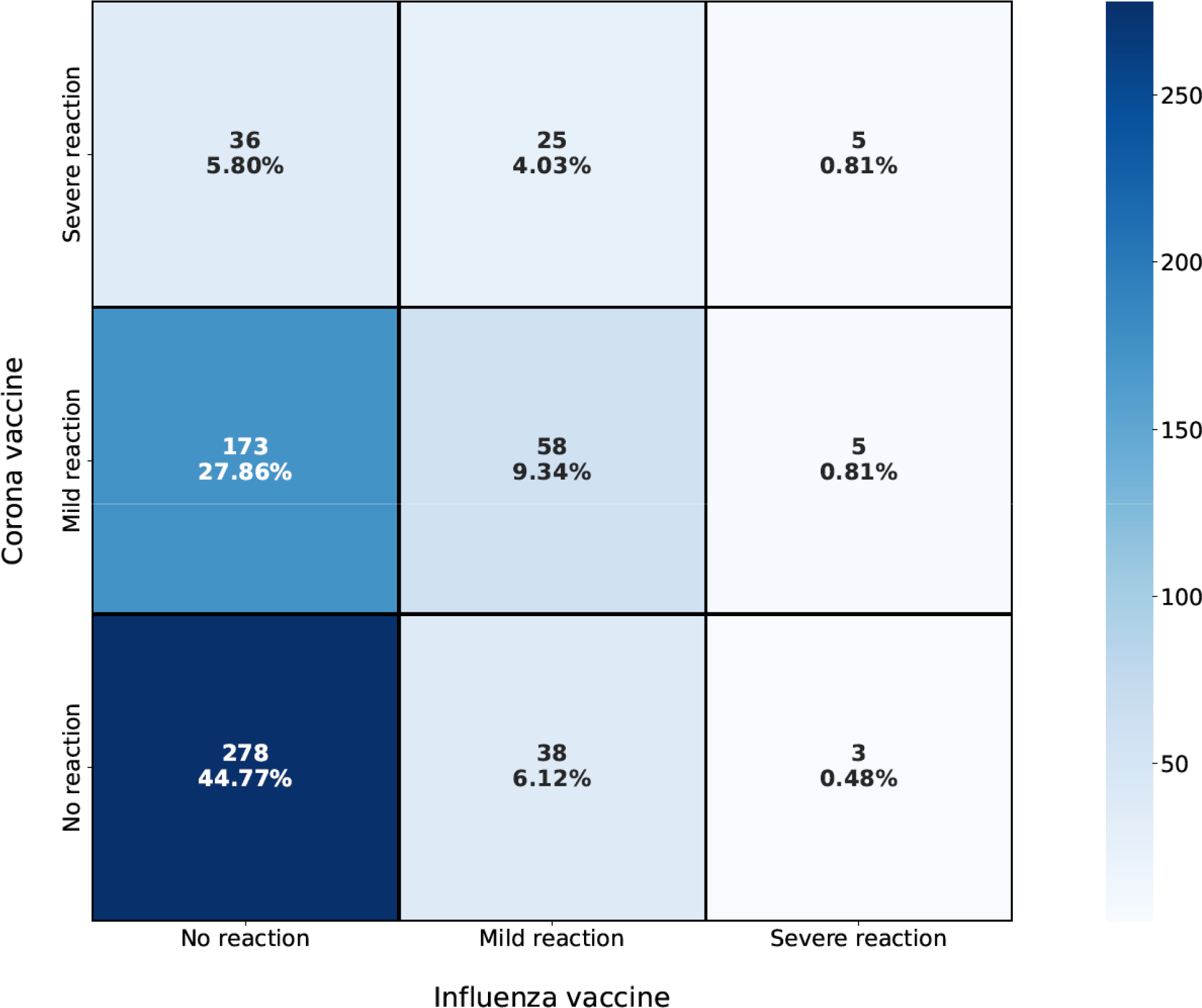
Comparison of self-reported reaction severity for COVID-19 vaccination and influenza vaccinatio among 621 individuals receiving both vaccines: number and percent of individuals reporting various combinations of no reaction, mild reaction, and severe reaction to the COVID-19 vaccine and influenza vaccine

